# Navigational Bronchoscopy versus Computed Tomography-guided Transthoracic Needle Biopsy for the Diagnosis of Indeterminate Lung Nodules: protocol and rationale for the VERITAS multicenter randomized trial

**DOI:** 10.1101/2023.11.22.23298915

**Authors:** Robert J. Lentz, Katherine Frederick-Dyer, Virginia B. Planz, Tatsuki Koyama, Matthew C. Aboudara, Briana Swanner, Lance Roller, See-Wei Low, Cristina Salmon, Sameer K. Avasarala, Todd C. Hoopman, Momen M. Wahidi, Kamran Mahmood, George Z. Cheng, James M. Katsis, Jonathan S. Kurman, Pierre-François D’Haese, Joyce Johnson, Eric L. Grogan, Charla Walston, Lonny Yarmus, Gerard A. Silvestri, Otis B. Rickman, Najib M. Rahman, Fabien Maldonado, the Interventional Pulmonary Outcomes Group (IPOG)

**Author notes:** <u>Correspondence to</u>: Fabien Maldonado, MD, Vanderbilt University Medical Center Division of Allergy, Pulmonary & Critical Care 1161 21st Avenue South, T-1218 MCN Nashville, TN 37232-2650.

## Abstract

**Background:** Lung nodule incidence is increasing. Many nodules require biopsy to discriminate between benign and malignant etiologies. The gold-standard for minimally invasive biopsy, computed tomography-guided transthoracic needle biopsy (CT-TTNB), has never been directly compared to navigational bronchoscopy, a modality which has recently seen rapid technological innovation and is associated with improving diagnostic yield and lower complication rate. Current estimates of the diagnostic utility of both modalities are based largely on non-comparative data with significant risk for selection, referral, and publication biases.

**Methods:** The VERITAS trial (na<u>V</u>igation <u>E</u>ndoscopy to <u>R</u>each Indeterminate lung nodules versus <u>T</u>ransthoracic needle <u>A</u>spiration, a randomized controlled <u>S</u>tudy) is a multicenter, 1:1 randomized, parallel-group trial designed to ascertain whether electromagnetic navigational bronchoscopy with integrated digital tomosynthesis is noninferior to CT-TTNB for the diagnosis of peripheral lung nodules 10-30 mm in diameter with pre-test probability of malignancy of at least 10%. The primary endpoint is diagnostic accuracy through 12 months follow-up. Secondary endpoints include diagnostic yield, complication rate, procedure duration, need for additional invasive diagnostic procedures, and radiation exposure.

**Discussion:** The results of this rigorously designed trial will provide high-quality data regarding the management of lung nodules, a common clinical entity which often represents the earliest and most treatable stage of lung cancer. Several design challenges are described. Notably, all nodules are centrally reviewed by an independent interventional pulmonology and radiology adjudication panel relying on pre-specified exclusions to ensure enrolled nodules are amenable to sampling by both modalities while simultaneously protecting against selection bias favoring either modality. Conservative diagnostic yield and accuracy definitions with pre-specified criteria for what non-malignant findings may be considered diagnostic were chosen to avoid inflation of estimates of diagnostic utility.

**Trial registration:** ClinicalTrials.gov NCT04250194

## INTRODUCTION

Indeterminate pulmonary nodules (IPNs) are increasingly discovered during lung cancer screening using low-dose computed tomography. Additionally, increased utilization of computed tomography (CT) for other indications has led to more frequent incidental identification of IPNs in the general population.^1^ Mechanisms to safely and accurately discriminate malignant from benign nodules will therefore be increasingly important.

The gold standard for minimally invasive lung nodule biopsy is CT-guided transthoracic needle biopsy (CT-TTNB), which is associated with diagnostic yield estimates around 90%, although studies focusing on smaller < 1.5 cm nodules have reported lower 70-80% yields.^2–4^ Complications occur in approximately 25% of cases, including pneumothorax in 15-25% with need for chest tube placement and hospital admission in 7% and significant hemorrhage in 1%.^5,6^ Navigational bronchoscopy (NB) is an alternative minimally invasive option with better safety profile, including pneumothorax in 1.6%, need for chest tube placement in 0.7%, and hemorrhage in 0.5%.^7^ Navigational bronchoscopy has the additional advantage of permitting contemporaneous sampling of multiple lung nodules and/or mediastinal and hilar lymph nodes in the same setting. However, diagnostic yield estimates have historically been lower than for CT-TTNB, ranging 38%-47% in registry studies to 73% in the largest prospective study to date.^7–9^ Most existing CT-TTNB and NB data consist of non-comparative studies (largely single center, retrospective, performed in high volume academic centers) at significant risk for selection, referral, and publication biases.^2,9^ These modalities have never been directly compared in a randomized trial despite both being considered standard of care for IPN biopsy.

A main limitation of NB compared to CT-TTNB has been an inability to confirm successful navigation to the lesion in real-time. The intraprocedural position of an IPN often differs from its expected location based on pre-procedure CT scan, an issue labeled CT-body divergence. Digital tomosynthesis allows CT-body divergence correction via intraprocedure 3D visualization of IPNs using standard 2D fluoroscopy. This technology has recently been integrated into the SuperDimension™ NB platform (Medtronic, Minneapolis, Minnesota, USA), with diagnostic yields around 80% achieved in recent studies, rivaling yields reported in the CT-TTNB literature.^10–13^.

We hypothesize that contemporary NB with intraprocedure digital tomosynthesis will have a non-inferior diagnostic accuracy for IPNs compared to CT-TTNB, with better safety profile. Herein we describe the methods, rationale, and potential implications of the findings of the VERITAS (naVigation Endoscopy to Reach Indeterminate lung nodules versus Transthoracic needle Aspiration, a randomized controlled Study) multicenter RCT designed to test this hypothesis.

## METHODS

### Trial Design

VERITAS is a multicenter, 1:1 randomized, noninferiority, parallel-group trial comparing gold-standard CT-TTNB to contemporary NB with respect to diagnostic utility for IPNs. This study was approved by the VUMC Institutional Review Board (IRB# 192142) and all local IRBs prior to enrollment at each site.

### Study Sites

This trial is being conducted at seven sites participating in the Interventional Pulmonary Outcomes Group, representing a mix of US tertiary care academic referral centers and private hospitals. Participating sites are listed in the clinicaltrials.gov registration (NCT04250194).

### Participants

#### Screening and eligibility

Adult patients with IPNs measuring 10-30 mm requiring tissue diagnosis (per referring specialist or lung nodule clinician) referred to lung nodule clinic, interventional pulmonology, or interventional radiology at participating centers are screened.

Eligibility criteria are listed in Table 1. Patients meeting inclusion criteria 1, 2a, and 2b and no exclusion criteria are approached for study inclusion. Radiographic (2c) and safety/feasibility (2d) inclusion criteria are centrally adjudicated after informed consent using de-identified CT images transmitted to the central coordinating center via the LungHive™ platform (Upstream Vision LLC, Nashville, TN, USA)^14^. Central radiographic adjudication ensures consistent segmentation of middle vs. outer third IPN location (an allocation stratification factor) and reliable exclusion of central IPNs less accessible to CT-TTNB. Safety/feasibility adjudication ensures all IPNs meet basic safety and accessibility criteria for both procedures with pre-specified reasons IPNs might be declined for biopsy by either modality in place, to preserve equipoise while limiting selection bias by proceduralists in either arm.

**Table 1.** Eligibility criteria.

|  |  |
| --- | --- |
| Inclusion criteria | 1. $\geq 18$ years of age at time of signing informed consent.<br>2. Referred for biopsy of a single IPN with the following characteristics:<br>a) Pre-test probability of malignancy of at least 10% using a validated clinical prediction model, which is either:<br>a. The Brock<sup>27</sup> model if no PET data are available, or<br>b. The Herder<sup>28</sup> model if PET data are available<br>b) Size between 10 and 30 mm (long diameter, inclusive).<br>c) *Peripheral in location, defined as occupying the middle or outer third lung zones, determined by segmentation using “CT Pulmo 3D” workflow (OsiriX, Pixmeo, Bernex, Switzerland).<sup>29</sup><br>d) *Technically amenable to both NB and CT-guided biopsy as confirmed by independent central adjudication panels with expertise in IPN biopsy, one comprised of interventional pulmonologists and one interventional radiologists, with the following pre-specified reasons for which a case can be deemed not technically amenable:<br>a. NB: no airway within 3 cm of the IPN<br>b. CT-TTNB: approach would require crossing a fissure or a bulla<br>Procedures may be deemed unsuitable for other reasons by either group with specific rationale recorded in the research record. |
| Exclusion criteria | 1. Central IPN that is accessible via endobronchial biopsy or linear endobronchial ultrasound-guided transbronchial needle aspiration.<br>2. Clinical indication for simultaneous biopsy of multiple IPNs.<br>3. Radiographically abnormal mediastinal or hilar lymph nodes for which |
|  | endobronchial ultrasound-guided sampling is clinically indicated.<br>4. Stereotactic body radiation therapy planned even if biopsies show no evidence of malignancy.<br>5. Contraindication to biopsy or deep sedation/general anesthesia.<br>6. Inability or unwillingness to comply with study follow-up schedule.<br>7. Previous randomization in this study.<br>8. Inability to provide documented informed consent.<br>9. Pregnant or nursing. |

\*Location in the middle or outer third and technical amenability to sampling via both modalities will be confirmed by central radiologic and interventional pulmonary and interventional radiology adjudication panels prior to randomization.

#### Discontinuation of allocated interventions

Patients may discontinue participation at any time with no further data recorded. Patients ineligible after central adjudication of inclusion criteria 2c and 2d are considered excluded prior to randomization. Additional exclusions include: new data available now meeting an exclusion criterion (e.g. positron emission tomography [PET] scan demonstrating nodal avidity), patients who decline the procedure or present for the procedure medically unstable (e.g. active chest pain, new arrhythmia) and are unable to reschedule.

#### Individuals performing the interventions

Participating bronchoscopists have performed more than 100 NB procedures in the prior two years, at least 25 with digital tomosynthesis. Participating interventional radiologists have performed more than 25 CT-TTNB procedures directed at peripheral lung lesions in the prior two years (in addition to regular experience performing CT-guided needle biopsy in other body locations).

#### Informed consent and Recruitment

Informed consent is obtained by investigators or research coordinators. Surrogate consent is not allowed. This study is being performed at high-volume centers and screens patients within typical referral pathways for both NB and CT-TTNB to increase the pool of prospective candidates and ensure a maximally generalizable sample is ultimately recruited. Effort is made to reschedule patients with scheduling issues or temporary bouts of medical instability.

### Allocation and Blinding

Patients are allocated 1:1 to NB or CT-TTNB. Randomization tables were computer-generated by the trial statistician then locked into the trial database preventing access by other personnel. Permuted blocks of 2 and 4 were utilized within the following strata: 1) nodule location: middle vs. outer third, 2) pre-test probability of malignancy per validated risk calculator: ≤50% vs. >50%, and 3) study site.

### Allocation is performed via a randomization module embedded in the research database

NB and CT-TTNB are performed in different departments using different layperson-identifiable technique preventing patient or proceduralist blinding. However, the central panel adjudicating diagnostic yield and accuracy (defined below), including pathologists performing central pathology review, will be blinded. No procedure for unblinding is required given the study design.

Study Procedures <u>Navigational bronchoscopy</u> A SuperDimension™ electromagnetic navigational bronchoscopy platform (Medtronic, Minneapolis, MN) with digital tomosynthesis capability (iLogic 7.2 or Illumisite™) is used for all NB procedures. NB procedure flow is detailed in Figure 1 with technical aspects reported in prior work.^10–13^ Navigation success is assessed using convex-probe radial endobronchial ultrasound (REBUS). If REBUS signature indicates no nodule or an eccentric nodule, digital tomosynthesis is performed to correct for CT-body divergence. Transbronchial needle aspiration (TBNA) is then performed with rapid on-site cytological evaluation in all cases to assess adequacy for immediate malignant diagnosis. Additional biopsy tools and techniques may then be used at the discretion of the proceduralist. Fluoroscopy is used to assess for pneumothorax with post-procedure thoracic ultrasound or formal chest radiography performed at the discretion of the proceduralist.

**Figure 1.**
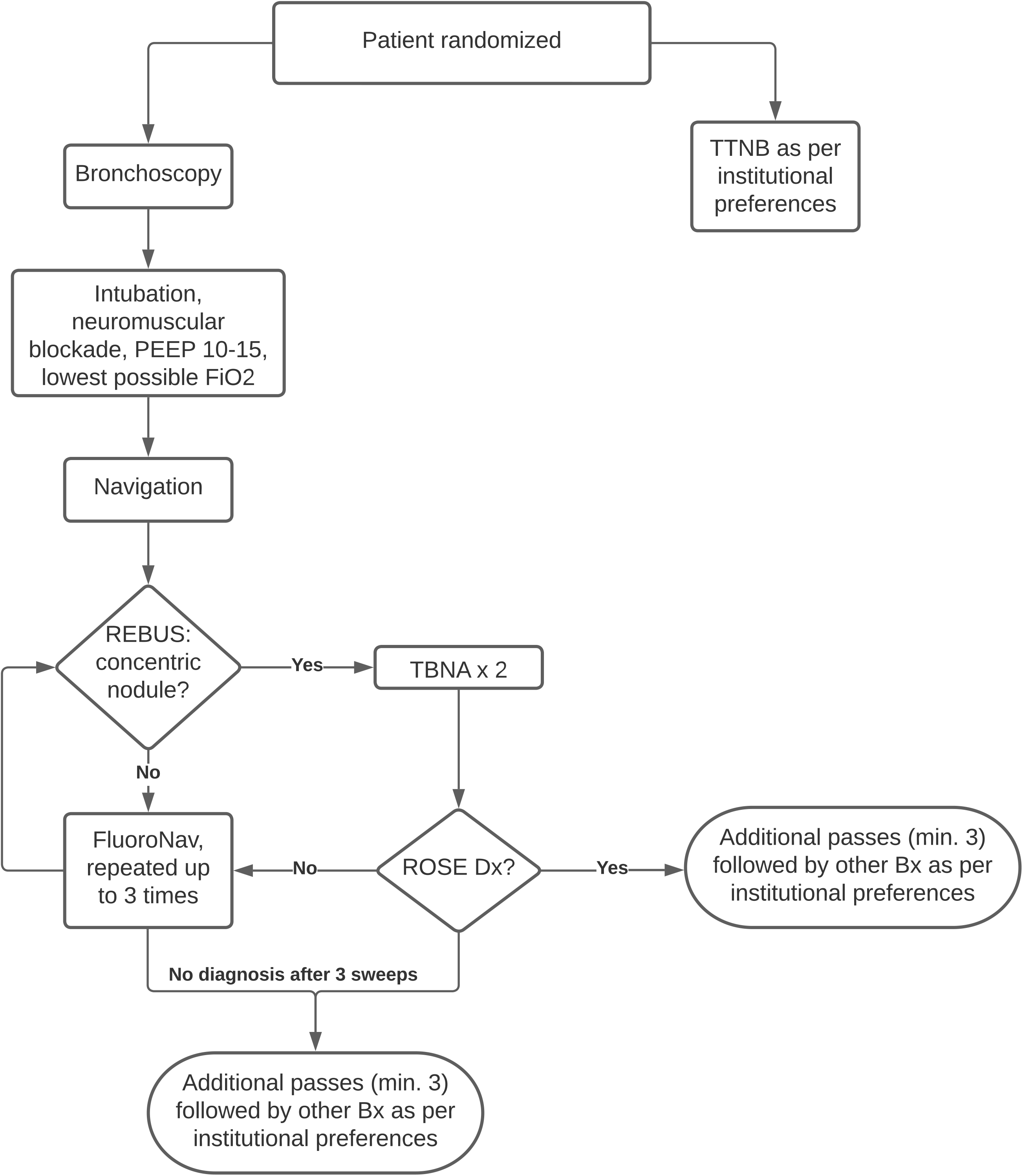
Biopsy Flow. PEEP = positive end-expiratory pressure. REBUS = concentric radial endobronchial ultrasound. TBNA = transbronchial needle aspiration. ROSE = rapid on-site cytological examination. Bx = biopsy.

### CT-guided transthoracic needle biopsy

Procedures are guided by a dedicated interventional CT scanner. Local anesthesia and moderate sedation or general anesthesia may be used according to local institutional preference. Biopsy number, core biopsy versus fine needle aspiration or both, use of coaxial needles, use of rapid on-site cytologic examination, and post-procedure examinations to assess for pneumothorax are at the discretion of the proceduralist per local institutional preferences.

### Concomitant and post-trial care

This protocol makes no prohibitions on patient care other than allocation to initial biopsy modality and all post-biopsy management is per managing physician(s), including need for additional biopsies after non-diagnostic study biopsies.

### Outcomes

Definitions of primary and secondary outcomes are detailed in Table 2. The primary outcome is diagnostic accuracy through 12 months (Figure 2). Secondary outcomes include diagnostic yield. Each biopsy result will be categorized as malignant, specific benign, or non-specific benign according to pre-specified criteria (Table 2). The diagnostic yield is conservative, with only biopsies with malignant or specific benign pathological findings considered diagnostic and eligible to be considered accurate at 12 month follow-up. All specimens not obviously malignant per local pathologist interpretation are reviewed by blinded central pathologists and all diagnostic categorizations will be agreed upon by a committee of blinded central lung nodule physicians based on these blinded central pathology reports.

**Figure 2.**
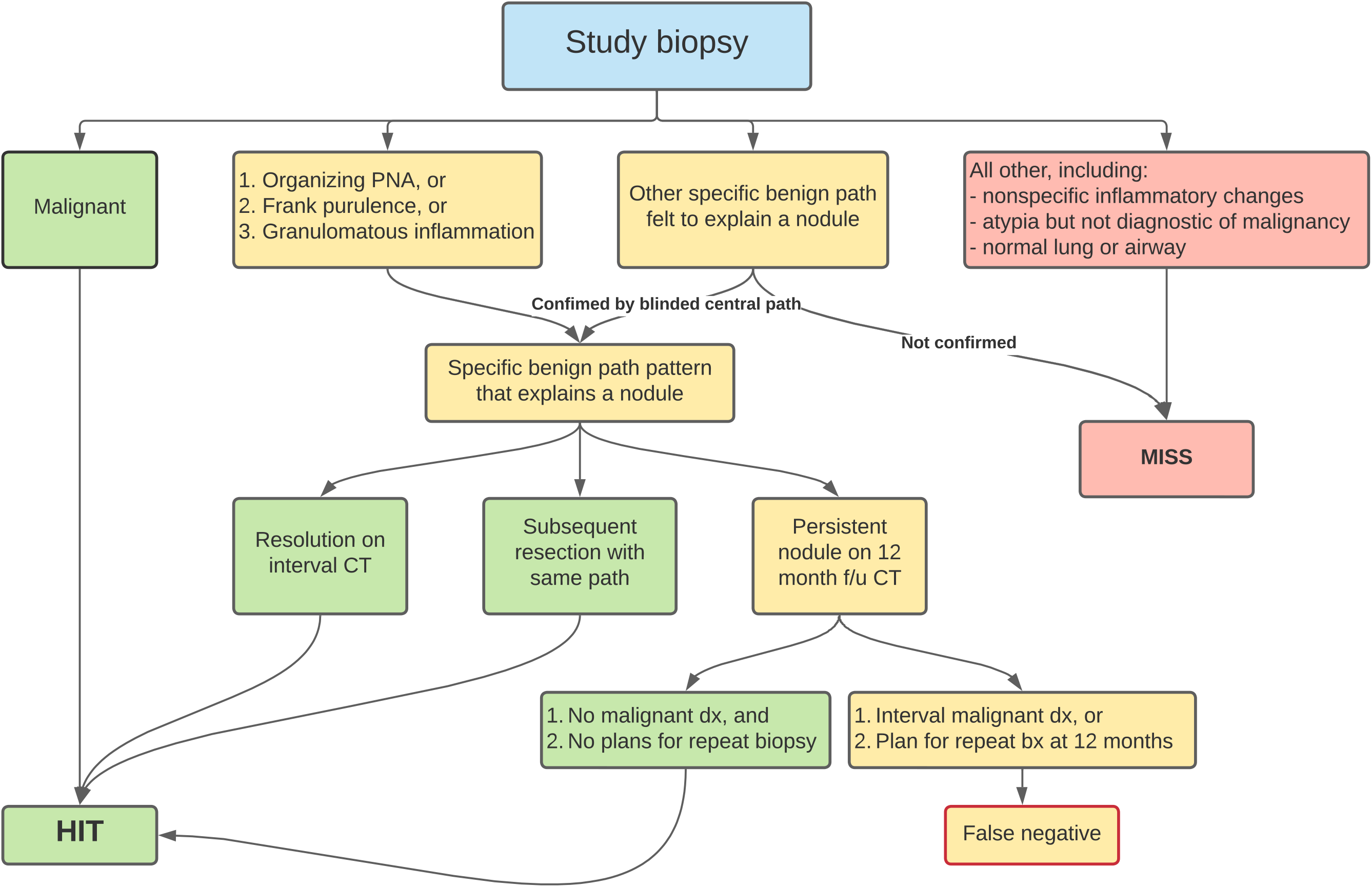
Diagnostic accuracy schematic.

**Table 2.**
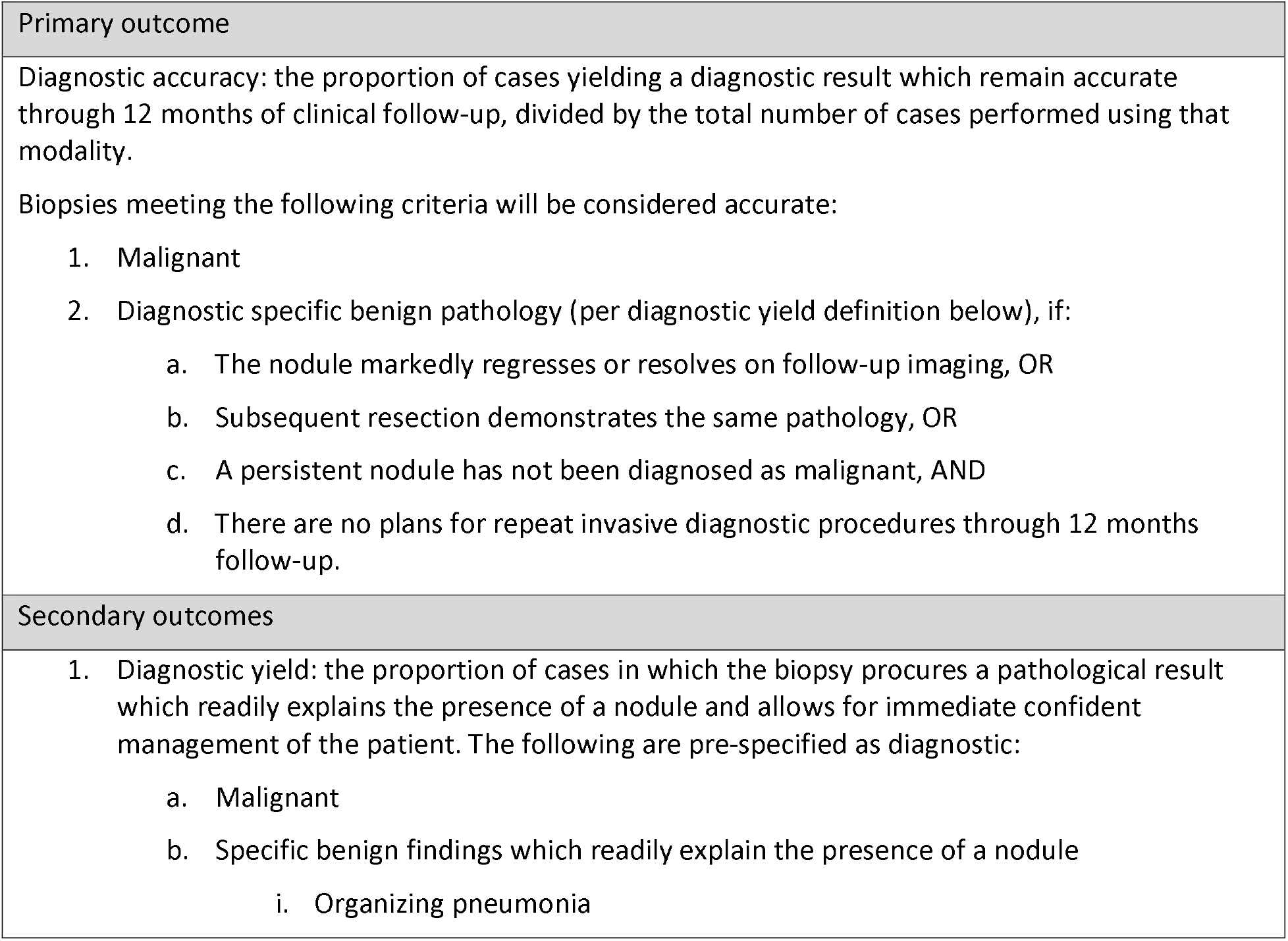
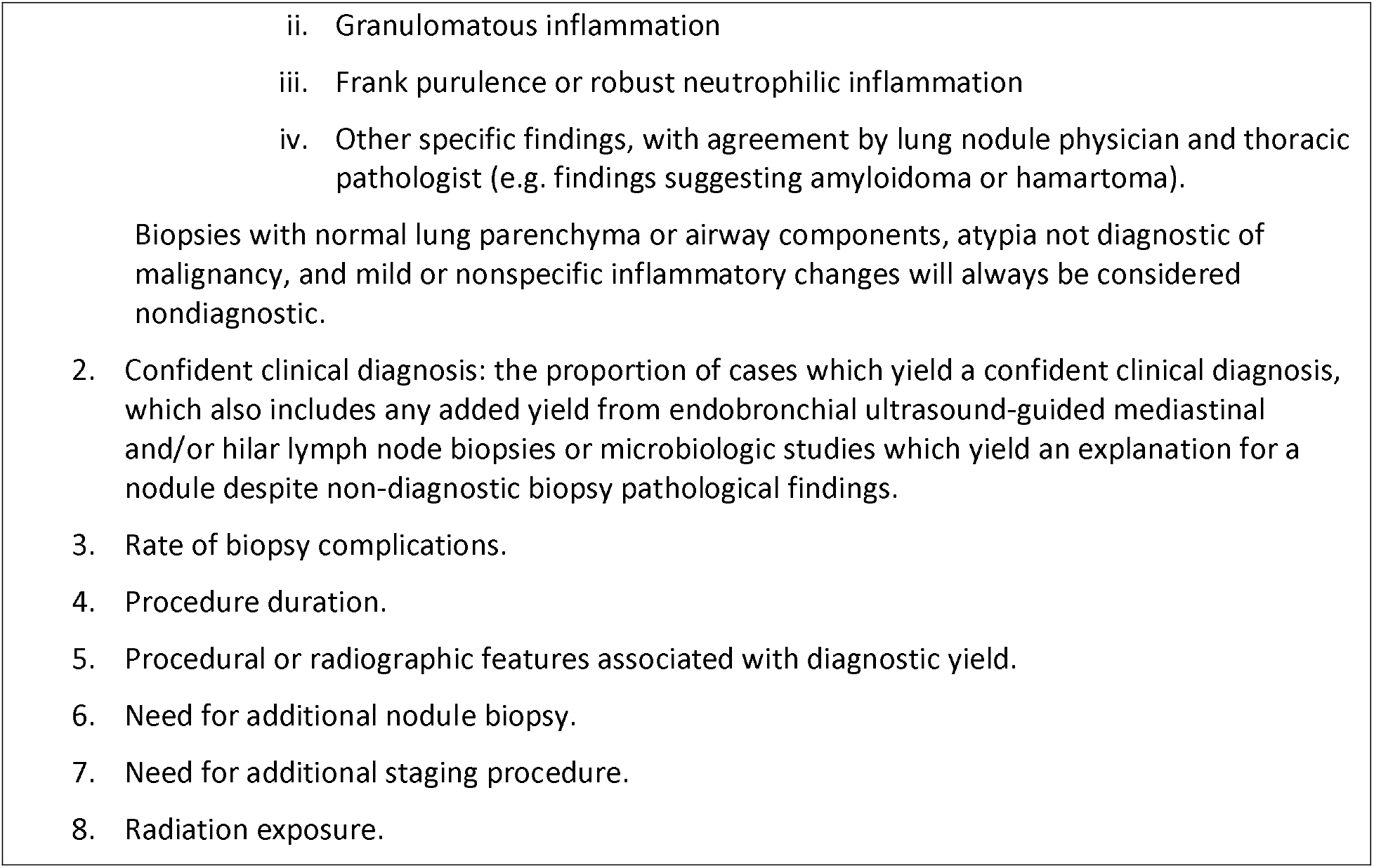
Trial outcomes.

Nodules are occasionally significantly smaller on day-of-procedure cross-sectional imaging, such as the scout CT scan just prior to CT-TTNB or same-day CT chest obtained for navigational bronchoscopy planning in patients with existing scans not compatible with navigation planning software, which causes the procedure to be aborted given the high likelihood of a benign nodule. These cases are considered diagnostic, as imaging integral to the allocated modality provided a result with high specificity for benign IPN, but follow-up diagnostic CT scan(s) to establish diagnostic accuracy as described in the primary outcome description are required. Any procedure declined by a proceduralist after allocation for reasons other than significant regression on day-of pre-procedure CT scan or those defined in “Discontinuation of allocated interventions” are considered nondiagnostic. Procedures started but not completed due to intraprocedural equipment malfunction, complication, patient intolerance, or other reasons are considered nondiagnostic. A procedure is “started” at time of induction of general anesthesia or initial insertion of a transthoracic needle.

The study schedule is detailed in Table 3. Patients with nodules determined to be malignant by trial biopsy are referred to appropriate oncological treatment. Nodules not definitively malignant by trial biopsy are subjected to standard clinical follow-up directed by managing physicians until follow-up CT imaging at least 12 months post-trial biopsy has been performed, unless a nodule resolves before 12 months.

**Table 3.** Study schedule.

| Study procedures | Visit 1 | Allocation | Visit 2<br>(biopsy) | Follow-Up<sup>a</sup><br>(after Visit 2 / biopsy) |  |  |  |
| --- | --- | --- | --- | --- | --- | --- | --- |
|  |  |  |  | 7 days<br>(+0-3d) | 3 months<br>(±14d) | 6 months<br>(±30d) | 12 months<br>(±30d) |
| Informed consent | X |  |  |  |  |  |  |
| Inclusion/exclusion criteria | X |  |  |  |  |  |  |
| Demographics | SOC |  |  |  |  |  |  |
| Medical history | SOC |  |  |  |  |  |  |
| Adverse events |  |  | X | X<sup>b</sup> | SOC | SOC | SOC |
| Physical exam | SOC |  |  |  | SOC | SOC | SOC |
| Labs | SOC |  |  |  |  |  |  |
| Pregnancy test |  |  | SOC |  |  |  |  |
| Radiology review | X |  |  |  |  |  |  |
| Pathology review |  |  |  | X | SOC<sup>c</sup> |  |  |
| CT scan (chest) | SOC |  | SOC |  | SOC |  |  |
| Allocation |  | X |  |  |  |  |  |
| NB or CT-TTNB (per allocation) |  |  | X |  |  |  |  |

Events marked with an 'X' will be performed per study protocol. Events marked 'SOC' will be performed per standard or usual care at the discretion of managing physicians.
<sup>a</sup>Follow-up cadence of 3, 6, and 12 months are not prescribed by the protocol as all events after intervention (biopsy) are per standard/usual care according to treating physicians. As 3, 6, and 12 months are common intervals for follow-up CT scans, medical records will be assessed at these intervals to determine if new results are available pertinent to trial outcomes.
<sup>b</sup>Patients will receive at least one follow-up contact by telephone or clinic visit (or by chart review in the event of patient incapacitation) 7-10 days after study biopsy.
<sup>c</sup>If subsequent biopsies or resection occurs.

### Statistical Methods

#### Sample size

A Frequentist approach was used to conduct an initial power calculation. Diagnostic accuracy of CT-TTNB was assessed at 90%, with noninferiority margin of 10%, one-sided type I error rate of 5%, and power of 80%, yielding a sample of n=112 per group, increased 15% to account for attrition to total n=258. The noninferiority margin was chosen in accordance with contemporary NB data utilizing digital tomosynthesis for CT-body divergence correction with diagnostic accuracy reports exceeding 80%. A diminution in diagnostic yield by less than 10% additionally seems clinically relevant as it is balanced by the assumed greater than 20% increase in pneumothorax risk in CT-TTNB versus NB. A simulation study confirmed the above operating characterizes remained accurate within a Bayesian paradigm in which there is no type I error rate (Statistical Analysis Plan, supplement).

#### Primary analysis

The primary analysis will use a Bayesian approach and be conducted on the per-protocol population, which may yield a more conservative estimate for the noninferiority objective than the intention-to-treat population.^15^ The probability of successful diagnostic accuracy for each treatment arm will be modeled with a Beta-Binomial distribution. A non-informative prior Beta (1,1) will be used for both treatment arms. We will conclude noninferiority of NB compared to CT-TTNB if the estimated probability that NB is worse than CT-TTNB by more than 10 percentage points is lower than 5% using the final posterior distributions of each trial group. The median estimate and 95% credible interval will be given for the diagnostic yield for NB and for CT-TTNB, as well as their difference. Superiority of NB will be concluded if the estimated probability that NB is better than CT-TTNB is higher than 95%.

#### Pre-hoc secondary and subgroup analyses

An intention-to-treat analysis of the primary outcome will also be performed including every randomized subject with diagnostic failure assigned to subjects who did not undergo study procedure.^16^ Pre-hoc analyses of subgroups will include the groups defined by two of the stratification factors (location in middle vs. outer third and pre-test probability ≤50% vs. >50%), nodule size, and presence vs. absence of a bronchus sign. Logistic regression models with interaction with treatment group will be fit to allow for comparison of diagnostic accuracy and yield differences across subgroups. An additional analysis including only those patients who underwent nodule biopsy (excluding those in which no biopsy was performed due to regression on same-day imaging or due to complication or equipment malfunction before biopsy procedures were complete) will also be performed.

#### Missing data and protocol nonadherence

The primary per-protocol analysis will include only those patients with complete follow-up data, which is conservative for the non-inferiority objective. We have planned adequate additional accrual to account for attrition with respect to the primary outcome and will monitor attrition rates as the trial progresses to determine if target accrual requires adjustment. The Bayesian design will allow for sample size change without inflation of the probability of incorrect conclusions. Sensitivity analyses will be performed for the primary per-protocol analysis in which all non-malignant diagnostic pathology results in patients lost to follow-up are considered true-negative and another in which they are all considered false-negative to determine the range of potential variation due to missing data.

We expect few instances of protocol non-adherence given the sole study intervention is allocation to NB vs. CT-TTNB with nearly all discrete procedural elements left up to experienced proceduralists.

#### Interim analysis

No interim analysis was performed as the primary outcome of diagnostic accuracy requires up to 12 months of radiographic follow-up; therefore, an analysis of this outcome part way through accrual could not be performed until one year later. Interim analyses for safety endpoints are felt unnecessary as both biopsy techniques are part of long-standing usual care.

### Data Collection and Monitoring

The central coordinating center for this trial is composed of the Principal Investigator (FM), central investigators (RJL, OBR), central research coordinators, interventional pulmonary and interventional radiology adjudicating panels, and study statistician (TK), with support from the Vanderbilt-Ingram Cancer Center Clinical Trials Office for independent logistical, monitoring, auditing, and regulatory oversight. No data monitoring committee was utilized as both procedures are current usual care.

All data are entered and maintained in a secured web-based Research Electronic Data Capture (REDCap) database by central and local investigators and research coordinators.^17,18^ Study personnel only have access to data entry forms pertinent to them. Source data verification is performed during standard audits. Reminders are sent to study physicians involved in the clinical follow-up of patients to increase the likelihood that 12-month surveillance CT data is available when necessary.

Both NB and CT-TTNB are associated with known clinical risks, including pneumothorax, airway hemorrhage, and respiratory failure. Efforts are made on post-procedure day 7-10 to assess for complications which have not already risen to investigator awareness. Adverse events are graded according to the NCI’s Common Terminology Criteria for Adverse Events, Version 5.0, dated November 27, 2017.

Patient confidentiality is protected by all study personnel as required by applicable laws and regulations with patient data transmitted to the central coordinating center only after full research informed consent has been obtained. All data is maintained in the aforementioned secured, HIPAA-compliant REDCap database. Protocol changes are submitted to all participating institution IRBs and not implemented at a given site until all approvals have been obtained.

### Trial status

The current protocol version is “23MAR2023” which corresponds with its date. Recruitment began 9/15/2020. The COVID-19 pandemic delayed accrual as both study procedures were considered aerosol-generating with risk to staff and therefore highly scrutinized early in the pandemic, and slow IRB and legal proceedings for non-COVID research delayed ethical approvals and data use agreements, hindering sub-site openings. The first sub-site opened 8/2021 with five additional opening through 12/2022. The final study procedure is expected to be performed in the Summer of 2023 with complete primary outcome accuracy data available 12 months later in Summer 2024.

A publication plan consistent with the International Committee of Medical Journal Editors will be created prior to analysis and publication of any data. Publication of results will be determined by the investigators, without limitations from the funding source. Reasonable written requests for the de-identified participant-level dataset and statistical code will be granted after publication of the primary manuscript. The full protocol will be made public via inclusion as a supplement to this manuscript.

## DISCUSSION

Indeterminate pulmonary nodules represent a common but unsettling clinical dichotomy: the vast majority are benign and clinically inconsequential while a minority are malignant and therefore extremely consequential. High-quality data regarding IPN biopsy methodologies are urgently needed to guide optimal clinical practice. The current IPN literature lacks methodological vigor in several domains, including dearth of randomized trials comparing biopsy modalities, overreliance on study designs highly prone to several biases, and imprecise definitions of biopsy diagnostic utility. The VERITAS trial was designed to counter these trends.

There are relatively few randomized trials of lung nodule biopsy. Most compare minor variations within NB or CT-TTNB rather than comparison across modalities^19–22^. Two randomized studies exist comparing a bronchoscopic modality to CT-TTNB: a 1998 study involving non-navigational bronchoscopic biopsy and a 2018 study involving REBUS-guided bronchoscopy. Both involved relatively large lung lesions and were underpowered for diagnostic utility (with n=30 and 50, respectively) with no difference found between groups in either.^23,24^ In lieu of high quality comparative data, the IPN biopsy literature is largely comprised of uncontrolled single-center studies highly prone to selection, referral, and publication biases, which are likely to have inflated diagnostic yield estimates and suppressed complication rates^9^.

The VERITAS trial is the first methodically robust study evaluating the respective diagnostic utility of contemporary NB and CT-TTNB. A rigorous determination of the diagnostic noninferiority of NB compared to CT-TTNB (if that is the ultimate finding) would have substantial implications, as the platform of bronchoscopy has several notable advantages: lower complication rate, simultaneous staging of the mediastinum using linear endobronchial ultrasound-guided (EBUS)-TBNA, and ability to biopsy multiple lung nodules in a single procedure. Therefore, results from this trial could directly affect mortality, morbidity, oncologic decision-making, and healthcare costs.

Generating accurate and generalizable results from this trial required careful design considerations to ensure neither modality has inherent advantages over the other regarding nodule eligibility. First, IPNs are screened via referrals in both interventional pulmonology and interventional radiology referral pathways to promote recruitment and ensure IPNs felt suitable for both procedures by referring clinicians are well represented. Second, all nodules are reviewed by a central adjudication panel in which an independent interventional pulmonologist and an interventional radiologists not involved in the procedure under consideration determine whether a nodule is amenable to each biopsy modality. Ineligibility determinations are based on conservative pre-specified criteria, which will serve to make these study results maximally generalizable. Likewise, lesions accessible via straightforward and high-yield bronchoscopic techniques, such as endobronchial biopsy or EBUS-TBNA, are excluded. Additional measures to avoid bias include meticulous screening to ensure a comprehensive CONSORT diagram.

We also carefully considered the definitions of “accurate” and “diagnostic” biopsies on which our main outcomes are predicated. Definitions of diagnostic utility vary widely in existing lung nodule diagnostics literature, predominantly related to how non-malignant results are classified. Vachani *et al* recently demonstrated in a simulation study that differences in how non-malignant biopsies are classified are expected to vary estimates of diagnostic yield as much as 20%.^25,26^ We chose conservative definitions of accuracy and yield, in which only non-malignant nodules demonstrating specific benign pathological findings (suggesting the nodule was truly accurately sampled during the procedure) will be considered diagnostic and eligible to be considered accurate if benign behavior is confirmed through 12 months of clinical follow-up.

In conclusion, the VERITAS trial will provide the first methodologically robust comparison of the diagnostic utility of contemporary NB versus gold standard CT-TTNB for IPNs. Trial design required careful attention to threats to validity and generalizability which have marred most existing lung nodule diagnostic literature. This trial may have major implications for the future of IPN diagnosis.

## Supporting information

Informed Consent Document

Statistical Analysis Plan

SPIRIT checklist

## Abbreviations

CT-TTNB: computed tomography-guided transthoracic needle biopsy

IPN: indeterminate pulmonary nodules

CT: computed tomography

NB: navigational bronchoscopy

RCT: randomized controlled trial

IRB: institutional review board

PET: positron emission tomography

REBUS: (convex probe) radial endobronchial ultrasound

TBNA: transbronchial needle aspiration

REDCap: Research Electronic Data Capture

EBUS: (linear) endobronchial ultrasound

## Data Availability

This is a description of a trial protocol - no data is reported

## ACKNOWLEDGEMENTS

Guarantor statement: The study sponsor-investigator, Fabien Maldonado, has full authority over study design; data collection, management, analysis and interpretation; writing of the report; and the decision to submit the report for publication. Study oversight was performed by the Vanderbilt Ingram Cancer Center, Clinical Trials Office, Coordinating Center. The funder had no role in study design, data collection/management/analysis/interpretation, oversight, writing of the report, or the decision to submit the report for publication.

## Summary COI statement

FM reports consulting fees from Intuitive. OBR reports consulting fees from Medtronic.

LY reports educational grants from Medtronic and consulting fees, educational and research grants from Olympus, Intuitive, and Johnson & Johnson.

MCA reports speaking fees from Medtronic and consulting fees from Noah Medical.

PFD is the CEO of Upstream Vision LLC, proprietor of LungHive software used in this work.

No other authors report any financial or non-financial conflicts of interest pertinent to this work.

## Author contributions

Study concept and design: FM, RJL, OBR, KFD, VP, TK, EG, LY, GS, NR

Acquisition of data: RJ, KFD, VP, MCA, BS, LR, SWL, CS, SKA, TCH, MW, KM, GC, JMK, JSK, JJ, PFD, CW, OBR, FM

Analysis and interpretation of data: n/a Drafting of the manuscript: RJL, FM, TK

All authors participated in critical revision of the manuscript for important intellectual content and provided final approval to submit this version of the manuscript and have agreed to be accountable for all aspects of the work.

