## Supplementary material for "Navigational Bronchoscopy versus Computed Tomography-guided Transthoracic Needle Biopsy for the Diagnosis of Indeterminate Lung Nodules: protocol and rationale for the VERITAS multicenter randomized trial": Informed Consent Document

VUMC Institutional Review Board
Informed Consent Document for Research

**Study Title:** Navigation endoscopy to reach indeterminate lung nodules versus transthoracic needle aspiration, a randomized controlled study (VERITAS)
**Version Date:** 03MAY2022
**PI:** Fabien Maldonado, MD

Name of participant: _______________________________________ Age: __________

**The following is given to you to tell you about this research study. Please read this form with care and ask any questions you may have about this study. Your questions will be answered. Also, you will be given a copy of this consent form.**

Concise Summary of Key Information
The purpose of this research study is to determine which procedure is the best for patients referred for biopsy of a lung nodule (growth in the lung) meeting the size and location requirements of the protocol. Two different procedures are available for lung nodule biopsy:

1) a computed tomography guided biopsy ("CT-guided biopsy") which consists of sampling the nodule from the "outside-in", through the chest wall with CT guidance, and
2) navigation bronchoscopy, which is a procedure using technology designed to guide a catheter through the natural airway route (wind-pipe and bronchi) to access the nodule.

There will be roughly 258 participants. Participants will be randomized (coin toss) to one group or the other and care will be considered standard of care, regardless of the group to which a participant is randomized. Participants will be followed as per standard of care and be in the study for up to one year.

The study does not present additional risks aside from the collection of personal information which will be deidentified and protected (see below).

If you are interested in learning more about this study, please continue to read below.

**What is the purpose of this study?**
You are being asked to take part in this research study because you were diagnosed with a lung nodule (growth in the lung ) for which a biopsy was recommended. Two different procedures are available for lung nodule biopsy: a computed tomography-guided biopsy ("CT-guided biopsy") which consists of sampling the nodule from the "outside-in", through the chest wall with CT guidance, and navigation bronchoscopy, which is a procedure using technology designed to guide a flexible bronchoscope (a small tube with a camera) through the natural airway route (wind-pipe and bronchi) to access the nodule. Both procedures have benefits and risk, and currently there is no consensus as to which procedure should be preferred in your situation. Both are outpatient procedures (same day).

CT-guided biopsies are performed by expert board-certified interventional radiologists in the radiology department. You will be lying down and after local anesthesia and conscious sedation (you will be sleepy, but not completely asleep), a biopsy needle will be introduced through the chest wall and directed towards the lung nodule. Confirmation of adequate position of the biopsy needle will be confirmed by CT imaging and biopsies obtained.

The accuracy of a CT-guided biopsy is overall 90%, and 75% for smaller nodules (less than 1.5 cm, or half an inch). Complications include lung collapse (when air leaks into the space between your lung and chest wall) in 15% of the cases, and hemorrhage (bleeding) in 1%.

Navigation bronchoscopy is performed by expert board-certified interventional pulmonologists in a bronchoscopy suite. You will be lying down and after general anesthesia (you will be fully asleep), a flexible bronchoscope (a small tube with a camera) will be introduced through the mouth and guided through the wind pipe and the bronchi towards the lung nodule. Confirmation of adequate position of the biopsy needle will be confirmed by ultrasound and X-ray and biopsies will be obtained.

The accuracy of navigation bronchoscopy is 75%, although recent data on current navigation bronchoscopy technology suggests an accuracy around 85%. Complications include lung collapse (when air leaks into the space between your lung and chest wall) in 2% of the cases and hemorrhage (bleeding) in 0.5%.

This study aims to determine which procedure is the best for patients in your situation. If you choose to be in this study, you will be randomized (coin toss) to one group or the other. We are trying to determine which procedure is the most accurate. You will not personally benefit from the study, but your care will be considered standard of care, regardless of the group to which you are randomized. As such, this study does not present additional risks for you aside from the collection of personal information which will be deidentified and protected (see below). You will be followed as per standard of care and be in the study for up to one year.

You do not have to be in this research study. You may choose not to be in this study and get other treatments without changing your healthcare, services, or other rights. You can stop being in this study at any time. If we learn something new that may affect the risks or benefits of this study, you will be told so that you can decide whether or not you still want to be in this study. Your medical record will contain a note saying you are in a research study and may contain some research information about you. Anyone you authorize to receive your medical record will also get this information.

**Side effects and risks that you can expect if you take part in this study:**
The side effects and risks that you can expect from participating in this study are the same you would be exposed to from the procedure outside of the study, namely:

<u>For bronchoscopy:</u>

*Common (>10%):* cough, sore throat, temporary hoarseness

*Uncommon (<10%):* lung collapse (when air leaks into the space between your lung and chest wall) requiring chest tube and admission to the hospital

*Rare (<1%):* bleeding, respiratory distress, infection, trauma to the airway or perforation, adverse reaction to medications used during the procedure, and extremely rarely (1/10,000) death.

For CT-guided biopsy:

*Common (>10%):* lung collapse (when air leaks into the space between your lung and chest wall), that does not require chest tube placement, chest wall pain, cough

*Uncommon (<10%):* lung collapse (when air leaks into the space between your lung and chest wall) requiring chest tube and admission to the hospital, bleeding

*Rare (<1%):* bleeding requiring blood transfusion, respiratory distress, infection, significant pain/discomfort, **vasovagal reaction (fainting associated with blood pressure changes), adverse reaction to medications used during the procedure,** and extremely rarely (1/10,000) death.

Non-physical risks of participating in the study: include the potential loss of confidentiality.

**Risks that are not known:**
Because both procedures are routinely performed in your situation, the likelihood of risks that we do not know about at this time is low.

**Good effects that might result from this study:**
The benefits to science and humankind that might result from this study: the information learned from this study will help future patients and physicians make more informed decisions about this common situation for which no high-quality comparative evidence is currently available. This study could lead to significant decrease in complications from these procedures and better patient care.

**Procedures to be followed:**
If you decide to participate in this study, you will be assigned randomly to one procedure or the other. There is currently no preferred procedure as both have advantages and disadvantages. Your care will accordingly be the same other patients would receive in the same situation. You will be asked to help us fill out forms to record information about the study and will be followed for up to one year after biopsy, although we may need to continue to follow you beyond one year if your medical care requires it.

**Payments for your time spent taking part in this study or expenses:**
You will not receive any payment as part of this study.

**Costs to you if you take part in this study:**

If you agree to take part in this research study, you and/or your insurance will not have to pay for the tests and treatments that are being done only for research. However, you are still responsible for paying for the usual care you would normally receive for the treatment of your illness. This includes treatments and tests you would need even if you were not in this study. These costs will be billed to you and/or your insurance.

You have the right to ask what it may cost you to take part in this study. If you would like assistance, financial counseling is available through the Vanderbilt Financial Assistance Program. The study staff can help you contact this program. You have the right to contact your insurance company to discuss the costs of your routine care (non-research) further before choosing to be in the study. You may choose not to be in this study if your insurance does not pay for your routine care (non-research) costs and your doctor will discuss other treatment plans with you.

**Payment in case you are injured because of this research study:**
If it is determined by Vanderbilt and the Investigator that an injury occurred as a direct result of the tests or treatments that are done for research, then you and/or your insurance will not have to pay for the cost of immediate medical care provided **at Vanderbilt** to treat the injury.

There are no plans for Vanderbilt to pay for any injury caused by the usual care you would normally receive for treating your illness or the costs of any additional care. There are no plans for Vanderbilt to give you money for the injury.

**Who to call for any questions or in case you are injured:**
If you should have any questions about this research study or if you feel you have been hurt by being a part of this study, please feel free to contact Dr. Fabien Maldonado at (615) 322-5000. If you cannot reach the research staff, please page the study doctor at (615) 875-8278

For additional information about giving consent or your rights as a person in this study, to discuss problems, concerns, and questions, or to offer input, please feel free to call the VUMC Institutional Review Board Office at (615) 322-2918 or toll free at (866) 224-8273.

**Reasons why the study doctor may take you out of this study:**
You may be taken out of the study if:

- Staying in the study would be harmful
- The study doctor believes it is best for you to be out of the study
- You do not follow directions and requirements for the study
- You become pregnant
- The study is cancelled
- You have a new injury or illness
- There may be other reasons to take you out of the study that we do not know at this time

**What will happen if you decide to stop being in this study?**
If you decide to stop being part of the study, you should tell your study doctor. Deciding to not be part of the study will not change your regular medical care in any way.

**Clinical Trials Registry:**
A description of this clinical trial will be available on www.clinicaltrials.gov, as required by U.S. Law. This website will not include information that can identify you. At most, the website will include a summary of the results. You can search this website at any time.

**Clinical Trials Reporting Program:**
Vanderbilt's NCI-Designated Cancer Center or the Sponsor registers National Cancer Institute (NCI)-supported clinical trials with NCI though their Clinical Trials Reporting Program (CTRP) to provide study related information. The data provided will include the following identifiable information that may identify you: birth month/year and five-digit zip code. NCI uses the data to manage and enhance the nation's investment in cancer research.

**Confidentiality:**
All efforts, within reason, will be made to keep your personal information in your research record confidential but total confidentiality cannot be guaranteed. Your information will be stored on password protected Vanderbilt computers and protected databases. Your data will be coded so they do not contain your name.
The Vanderbilt University Office of Research will be used as a central location for data processing and management. REDCap (Research Electronic Data Capture) is a secure, web-based application that is the database. REDCap servers are housed in a local data center at Vanderbilt, and all web-based information transmission is encrypted. REDCap was developed specifically around HIPAA-Security guidelines. Only those involved with this study will have access to your information. Any information that is shared or published will not include identifiable information. Your information will be kept indefinitely. Vanderbilt may share your information, without identifiers, to others or use it for other research projects not listed in this form. Vanderbilt, Dr. Maldonado and his staff will comply with any and all laws regarding the privacy of such information. There are no plans to pay you for the use or transfer of this de-identified information.

This study may have some support from the National Institutes of Health (NIH). If so, your study information is protected by a Certificate of Confidentiality. This Certificate allows us, in some cases, to refuse to give out your information even if requested using legal means.

It does not protect information that we have to report by law, such as child abuse or some infectious diseases. The Certificate does not prevent us from disclosing your information if we learn of possible harm to yourself or others, or if you need medical help.

Study Title: Navigation endoscopy to reach indeterminate lung nodules versus transthoracic needle aspiration, a randomized controlled study (VERITAS)
Version Date: 03MAY2022
PI: Fabien Maldonado, MD

---

Disclosures that you consent to in this document are not protected. This includes putting research data in the medical record or sharing research data for this study or future research. Disclosures that you make yourself are also not protected.

**Privacy:**
Any samples and information about you may be made available to others to use for research. To protect your privacy, we will not release your name. You will not receive any benefit as a result of the tests done on your samples. These tests may help us or other researchers learn more about the causes, risks, treatments, or how to prevent this and other health problems.

**Study Results:**
Once completed, the study results will be analyzed and presented to the research community as a scientific abstract and published manuscript which will be available to the public.

**Authorization to Use/Disclose Protected Health Information**

**What information is being collected, used, or shared?**
To do this research, we will need to collect, use, and share your private health information. By signing this document, you agree that your health care providers (including both Vanderbilt University Medical Center and others) may release your private health information to us, and that we may use any and all your information that the study team believes it needs to conduct the study. Your private information may include things learned from the procedures described in this consent form, as well as information from your medical record (which may include information such as HIV status, drug, alcohol or STD treatment, genetic test results, or mental health treatment).

**Who will see, use or share the information?**
The people who may request, receive or use your private health information include the study sponsor Vanderbilt Medical Center and its agents or contractors, study safety monitors and auditors, data managers and other agents and contractors used by the study team, researchers and study team members. Additionally, we may share your information with other people at Vanderbilt, for example if needed for your clinical care or study oversight. Also, your records may be seen by people from regulatory authorities (like the US Food and Drug Administration [FDA]), auditors, and the IRB. By signing this form, you give permission to the research team to share your information with others outside of Vanderbilt University Medical Center. This may include the funder of the study and its agents or contractors, outside providers, government agencies and other sites in the study. We try to make sure that everyone who sees your information keeps it confidential, but we cannot guarantee that your information will not be shared with others. If your information is disclosed by your health care providers or the research team to others, federal and state confidentiality laws may no longer protect it.

**Do you have to sign this Authorization?**
You do not have to sign this Authorization, but if you do not, you may not join the study.

VUMC Institutional Review Board
Informed Consent Document for Research

Study Title: Navigation endoscopy to reach indeterminate lung nodules versus transthoracic needle aspiration, a randomized controlled study (VERITAS)
Version Date: 03MAY2022
PI: Fabien Maldonado, MD

**How long will your information be used or shared?**
Your Authorization for the collection, use, and sharing of your information does not expire. Additionally, you agree that your information may be used for similar or related future research studies.

**What if you change your mind?**
You may change your mind and cancel this Authorization at any time. If you cancel, you must contact the Principal Investigator in writing to let them know by using the contact information provided in this consent form. Your cancellation will not affect information already collected in the study, or information that has already been shared with others before you cancelled your authorization.

If you decide not to take part in this research study, it will not affect your treatment, payment or enrollment in any health plans or affect your ability to get benefits. You will get a copy of this form after it is signed.

STATEMENT BY PERSON AGREEING TO BE IN THIS STUDY

I have read this consent form and the research study has been explained to me verbally. All my questions have been answered, and I freely and voluntarily choose to take part in this study.

________________ ________________________________
Date Signature of patient/volunteer

Consent obtained by:

________________ ________________________________
Date Signature

________________________________
Printed Name and Title

This box is for IRB USE ONLY Do not edit or delete
Date of IRB Approval: 09/20/2022
Date of Expiration: 09/19/2023
Institutional Review Board
VANDERBILT
