## Supplementary material for "Navigational Bronchoscopy versus Computed Tomography-guided Transthoracic Needle Biopsy for the Diagnosis of Indeterminate Lung Nodules: protocol and rationale for the VERITAS multicenter randomized trial": Statistical Analysis Plan

VERITAS

Statistical Analysis Plan

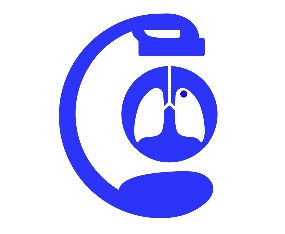

THO 19102: Na**V**igation **E**ndoscopy to **R**each **I**ndeterminate lung nodules versus **T**ransthoracic needle **A**spiration, a randomized controlled **S**tudy (VERITAS)

Principal Investigator:

Fabien Maldonado, MD

Professor of Medicine and Thoracic Surgery

Associate Professor of Mechanical Engineering

Vanderbilt University School of Medicine

**Sample Size**

A Frequentist approach was used to conduct an initial power calculation. Diagnostic accuracy of CT-TTNB was assessed at 90%, with noninferiority margin of 10%, one-sided type I error rate of 5%, and power of 80%, yielding a sample of n=112 per group, increased 15% to account for attrition to total n=258. The noninferiority margin was chosen in accordance with contemporary NB data utilizing digital tomosynthesis for CT-body divergence correction with diagnostic accuracy reports around 80%. A diminution in diagnostic yield by less than 10% additionally seems balanced by the greater than 20% increase in pneumothorax risk in CT-TTNB versus NB.

A simulation study confirmed the above operating characterizes remained accurate within a Bayesian paradigm in which there is no type I error rate. To generate data, we first postulated the true distribution of the diagnostic accuracy using a Beta distribution, which gives the specific mean and standard deviation. For example, Beta(141,35) is used to represent a distribution with mean 0.80 and standard deviation 0.03 and 0.01.

| Distribution | Mean | SD | Distribution | Mean | SD |
| --- | --- | --- | --- | --- | --- |
| Beta(159,106) | 0.6 | 0.03 | Beta(1439,960) | 0.6 | 0.01 |
| Beta(163, 70) | 0.7 | 0.03 | Beta(1469,630) | 0.7 | 0.01 |
| Beta(141,35) | 0.8 | 0.03 | Beta(1279,320) | 0.8 | 0.01 |
| Beta(89,10) | 0.9 | 0.03 | Beta(809,90) | 0.9 | 0.01 |

Based on simulation of 10,000 trials, we determined a final sample size of 112 per arm was sufficient to limit the probability of incorrect decisions to acceptable levels. Table S1 summarizes the results of the simulation study, in which we assume the true diagnostic yield is 60 to 90% for each approach. When the true state is inferiority such that the NB is worse than CT-TTNB by 10 percentage points, the probability to incorrectly conclude non-inferiority is around 5%. When the true diagnostic accuracy is equal under CT-TTNB and NB (non-inferiority), the probability of concluding NB’s superiority is slightly below 5%. Thus, n=112, coupled with our decision rule, will limit the probabilities of these errors (type-I-error equivalent) at acceptable levels. The noninferiority margin of 10 percentage-points was chosen in accordance with contemporary NB data utilizing digital tomosynthesis for CT-body divergence correction with diagnostic accuracy reports around 80%. A diminution in diagnostic yield by less than 10% additionally seems balanced by the greater than 20% increase in pneumothorax risk in CT-TTNB versus NB. Finally, accounting for anticipated attrition rate of 15%, a sample size of n=129 (258 total) was ascertained.

Table S1. Bayesian analysis operating characteristics based on simulation study assuming SD=0.03.

|  |  |  |  | Final analysis: probability of concluding… | |
| --- | --- | --- | --- | --- | --- |
| True TTNB  Mean | True NB  Mean | True State | Difference | NB is non-inferior | NB is superior |
| 0.7 | 0.6 | Inferiority | -0.1 | 4.9 | <1 |
| 0.8 | 0.7 | Inferiority | -0.1 | 5.0 | <1 |
| 0.9 | 0.8 | Inferiority | -0.1 | 5.2 | <1 |
| 0.8 | 0.6 | Inferiority | -0.2 | <1 | <1 |
| 0.9 | 0.7 | Inferiority | -0.2 | <1 | <1 |
| 0.7 | 0.7 | Non-inferiority | 0.0 | 45.1 | 4.9 |
| 0.8 | 0.8 | Non-inferiority | 0.0 | 54.3 | 5.1 |
| 0.9 | 0.9 | Non-inferiority | 0.0 | 75.4 | 4.8 |
| 0.6 | 0.7 | Superiority | 0.1 | 46.9 | 46.7 |
| 0.7 | 0.8 | Superiority | 0.1 | 44.3 | 52.8 |
| 0.8 | 0.9 | Superiority | 0.1 | 32.2 | 67.4 |
| 0.6 | 0.8 | Superiority | 0.2 | 4.8 | 95.2 |
| 0.7 | 0.9 | Superiority | 0.2 | 1.5 | 98.5 |

*Simulation study using SD = 0.01 yielded results not significantly different than those noted for SD = 0.03.

***Primary Analysis***

The primary analysis will use a Bayesian approach. This approach was chosen because it makes the analysis straightforward with simple-to-interpret results and allows for sample size adjustment without inflation of the probability of incorrect conclusions should attribution/loss to follow-up exceed that estimated for the planned sample size.

The primary analysis will be conducted on the per-protocol population, which may yield a more conservative estimate for the non-inferiority objective than the intention-to-treat population. The probability of successful diagnostic accuracy for each treatment arm will be modeled with a Beta-Binomial distribution. A non-informative prior Beta(1,1), will be used for both treatment arms. Analysis will be conducted when complete follow-up data from all randomized patients are available. The conclusion will be based on the final posterior distributions. Noninferiority of NB compared to CT-TNNB will be concluded if the estimated probability that NB is worse than CT-TTNB by more than 10 percentage points is lower than 5%. Similarly, superiority of NB will be concluded if the estimated probability that NB is better than CT-TTNB is higher than 95%. The median estimate and 95% credible interval will be given for the diagnostic yield for NB and for CT-TTNB based on their posterior distributions. Difference in diagnostic yields will be estimated by sampling from the posterior distributions.

A secondary intention-to-treat sensitivity analysis will be performed including every randomized subject, assigning diagnostic failure to subjects lost to follow-up without complete 12-month data.

***Interim analysis***

No interim analysis is planned for this trial. Interim analyses typically assess for futility and efficacy stops regarding the primary outcome part-way through a trial. However, the primary outcome of diagnostic accuracy requires up to 12 months of radiographic follow-up in patients not determined to have malignant nodules by study biopsy. Therefore, an analysis of this outcome halfway through accrual could not be performed until one year after that enrollment midpoint was reached, at which time we expect enrollment to be complete for the entire planned accrual. An interim analysis of the primary endpoint is therefore impracticable. Interim analyses for safety endpoints are often commonly performed in investigations of new devices or novel interventions, but as both study procedures represent long-standing standard of care options for the tissue sampling of lung nodules, complications of these procedures themselves are not considered risks unique to this research. The primary risk to participation in this research involves patient privacy and confidentiality, any breach of which will be reported to the PI and appropriate regulators on an ongoing basis as described later in this protocol, which does not require interim analysis.

**Pre-hoc secondary and subgroup analyses**

An intention-to-treat analysis of the primary outcome will also be performed, as suggested by CPMP guidelines (main manuscript reference #14), including every randomized subject with diagnostic failure assigned to subjects who did not undergo study procedure. Pre-hoc analyses of subgroups will include the groups defined by two of the stratification factors (location in middle vs. outer third and pre-test probability ≤50% vs. >50%), nodule size, and presence vs. absence of a bronchus sign. Logistic regression models with interaction with treatment group will be fit to allow for comparison of diagnostic accuracy and yield differences across subgroups. Same-day preprocedural imaging (e.g. scout CT scan just prior to CT-TTNB or CT chest for navigational bronchoscopy planning in patients with existing scans incompatible with navigational software) occasionally demonstrates regression or resolution of the target lesion. As these imaging studies are considered an integral part of these procedure and this result is highly predictive of benign IPNs, these procedures will be considered diagnostic for the primary analysis, but we will also report sensitivity analyses in which these procedures are considered (1) non-diagnostic, or (2) will be excluded.

**Missing data and sensitivity analyses**

The primary per-protocol analysis will include only those patients with complete follow-up data, which is conservative for the non-inferiority objective. We have planned adequate additional accrual to account for attrition with respect to the primary outcome and will monitor attrition rates as the trial progresses to determine if target accrual requires adjustment. The Bayesian design will allow for sample size change without inflation of the probability of incorrect conclusions. Sensitivity analyses will be performed for the primary per-protocol analysis in which all non-malignant diagnostic pathology results in patients lost to follow-up are considered true-negative and another in which they are all considered false-negative to determine the range of potential variation due to missing data.

**Variables recorded**

- Past medical history
- Demographic data
- Smoking history
- Current medications
- Chest CT characteristics:
  - Location of the nodule (lobe and segment)
  - Nodule size (longest axis)
  - Distance to the nearest visible bronchus on either CT plane (axial, coronal or sagittal)
  - Bronchus sign (airway aligned with nodule)
  - Distance to the nearest parietal pleura
- PET scan characteristics (FDG uptake (SUV))
- Procedural characteristics:
  - Duration of procedure
  - Number and type of biopsies
  - REBUS signatures after initial navigation and after digital tomosynthesis, if used
  - Use of digital tomosynthesis
  - Radiation exposure
  - Complications (pneumothorax, bleeding, any others)
- Time to procedure (delay between decision to biopsy and procedure)
- Procedure duration
- Need for another biopsy modality
- Need for an additional staging procedure
- Follow-up data

**Primary endpoint**

The primary endpoint will be diagnostic accuracy, defined as the proportion of cases yielding a diagnostic result which remain accurate through 12 months of clinical follow-up, divided by the total number of cases allocated to that modality, comparing the navigation bronchoscopy to the CT-guided biopsy pathway.

Biopsies meeting the following criteria will be considered accurate:

- Malignant
- Diagnostic specific benign pathology (per diagnostic yield definition below), if:
  - The nodule markedly regresses or resolves on follow-up imaging, OR
  - Subsequent resection demonstrates the same pathology, OR
  - A persistent nodule has not been diagnosed as malignant, AND

**Secondary endpoints**

1. Diagnostic yield: the proportion of cases in which the biopsy procures a pathological result which readily explains the presence of a nodule and allows for immediate confident management of the patient. The following are pre-specified as diagnostic:
   a. Malignant
   b. Specific benign findings which readily explain the presence of a nodule
      i. Organizing pneumonia
      ii. Granulomatous inflammation
      iii. Frank purulence or robust neutrophilic inflammation
      iv. Other specific findings, with agreement by lung nodule physician and thoracic pathologist (e.g. findings suggesting amyloidoma or hamartoma).

Biopsies with normal lung parenchyma or airway components, atypia not diagnostic of malignancy, and mild or nonspecific inflammatory changes will always be considered nondiagnostic.

2. Confident clinical diagnosis: the proportion of cases which yield a confident clinical diagnosis, which also includes any added yield from endobronchial ultrasound-guided mediastinal and/or hilar lymph node biopsies or microbiologic studies which yield an explanation for a nodule despite non-diagnostic biopsy pathological findings.
3. Rate of biopsy complications.
4. Procedure duration.
5. Procedural or radiographic features associated with diagnostic yield.
6. Need for additional nodule biopsy.
7. Need for additional staging procedure.
8. Radiation exposure.

All biopsies not diagnosed as malignant will be reviewed by a central expert lung pathologist blinded to the biopsy modality. After close to accrual, the reports generated by this blinded central pathologist will be reviewed by a panel of blinded expert lung nodule clinicians who will adjudicate diagnostic yield and accuracy outcomes using the definitions defined above.
